# Predicting future ocular *Chlamydia trachomatis* infection prevalence using serological, clinical, molecular, and geospatial data

**DOI:** 10.1101/2021.07.19.21260623

**Authors:** Christine Tedijanto, Solomon Aragie, Zerihun Tadesse, Mahteme Haile, Taye Zeru, Scott D. Nash, Dionna M. Wittberg, Sarah Gwyn, Diana L. Martin, Hugh J.W. Sturrock, Thomas M. Lietman, Jeremy D. Keenan, Benjamin F. Arnold

## Abstract

Trachoma is an infectious disease characterized by repeated exposures to *Chlamydia trachomatis* (*Ct*) that may ultimately lead to blindness. District-level estimates of clinical disease are currently used to guide control programs. However, clinical trachoma is a subjective indicator. Serological markers present an objective, scalable alternative for monitoring and targeting of more intensive control efforts. We hypothesized that IgG seroprevalence in combination with geospatial layers, machine learning, and model-based geostatistics would be able to accurately predict future community-level ocular *Ct* infections detected by PCR. Among 40 communities in the hyperendemic Amhara region of Ethiopia, median *Ct* infection prevalence among children 0-5 years old increased from 6% at enrollment to 29% at month 36. Seroprevalence was the strongest concurrent predictor of infection prevalence at month 36 among children 0-5 years old (cross-validated R^2^ = 0.75, 95% CI: 0.58-0.85), though predictive performance declined substantially with increasing temporal lag between predictor and outcome measurements. Geospatial variables, a spatial Gaussian process, and stacked ensemble machine learning did not meaningfully improve predictions. Serological markers among children 0-5 years old may be a promising programmatic tool for identifying communities with high levels of active ocular *Ct* infections, but accurate, future prediction in the context of changing transmission remains a challenge.

## INTRODUCTION

Trachoma, caused by ocular infection with the bacterium *Chlamydia trachomatis* (*Ct*), is a leading infectious cause of blindness worldwide (1) and has been targeted for elimination as a public health problem by 2030 (2). The World Health Organization’s SAFE strategy (Surgery, Antibiotics, Facial cleanliness, and Environmental improvement) has been successful in countries across Asia and the Middle East, achieving elimination as a public health problem in many cases (2). Yet, trachoma is a persistent challenge in pockets of Africa, including some areas of Ethiopia that remain hyperendemic despite over 10 years of control activities (3). The ability to efficiently identify potential areas of ongoing transmission for follow-up surveys and more intensive interventions is crucial for the trachoma endgame.

Trachoma elimination programs are currently guided by estimates of clinical disease markers, including trachomatous inflammation — follicular (TF), in evaluation units (EUs) of 100,000-250,000 people (4). Evidence of trachoma clusters at the village- or sub-village level throughout Africa (5–10) suggest that aggregate estimates may mask heterogeneity in infection: high-transmission villages may be missed by sampling design or their signal may be “washed out” in EU-level averages. Fine-scale estimates of trachoma could facilitate targeted allocation of limited resources to communities with the highest burden (11) and reduce unnecessary antibiotic use and subsequent selection for antibiotic resistance (12).

Mass drug administration (MDA) of azithromycin is currently recommended for EUs with TF prevalence above 5% among children aged 1-9 years old (2). Clinical disease states are relevant signals of progression towards conjunctival scarring and ultimately blindness (1) but are subject to misclassification, even by experienced graders (13). Immunoglobulin G (IgG) antibody responses to Pgp3 and CT694 antigens are a more objective alternative and have been identified as sensitive, specific, and durable indicators of past ocular *Ct* infection (14, 15). In addition, dried blood spot specimens used to assess serological markers are easy to collect, and *Ct* antigens can be included in multiplexed, integrated serosurveillance platforms to simultaneously and cost-effectively monitor numerous pathogens (16).

Thus far, efforts to predict future trachoma prevalence at the village and district level have had modest success (17, 18) but have not considered serology or recent advances in machine learning and geostatistics that may facilitate fine-scale prediction. We hypothesized that models incorporating trachoma indicators (clinical disease, ocular *Ct* infection identified by polymerase chain reaction (PCR), and IgG response to *Ct* antigens), remotely sensed geospatial layers, and spatial structure would accurately predict future community-level *Ct* infection prevalence. We also hypothesized that seroprevalence would be a more accurate and stable predictor of *Ct* infections compared to clinical disease and that communities with high levels of infection would be geographically clustered in stable foci of transmission (“hotspots”). We tested our hypotheses using data from the WASH Upgrades for Health in Amhara (WUHA) randomized controlled trial (<u>NCT02754583</u>) (19).

## RESULTS

### Study population and setting

WUHA was a randomized controlled trial designed to evaluate the impact of a comprehensive water, sanitation, and hygiene (WASH) intervention on ocular *Ct* infection. The trial was conducted in forty communities in a dry, mountainous region of the Wag Hemra Zone of Amhara, Ethiopia (**Figure 1**). MDA was conducted for seven consecutive years in the study communities before the study began but was suspended for the duration of the trial. Baseline measurements were collected in December 2015 (month 0), and follow-up visits occurred annually for three years thereafter (months 12, 24, and 36). Clinical, serological, and molecular trachoma indicators were measured among randomly sampled children ages 0-9 years old at each visit. Data were combined across the two intervention arms for this secondary analysis as no difference was observed for the primary endpoint of ocular *Ct* infection at the end of the study period [manuscript under review].

**Figure 1.**
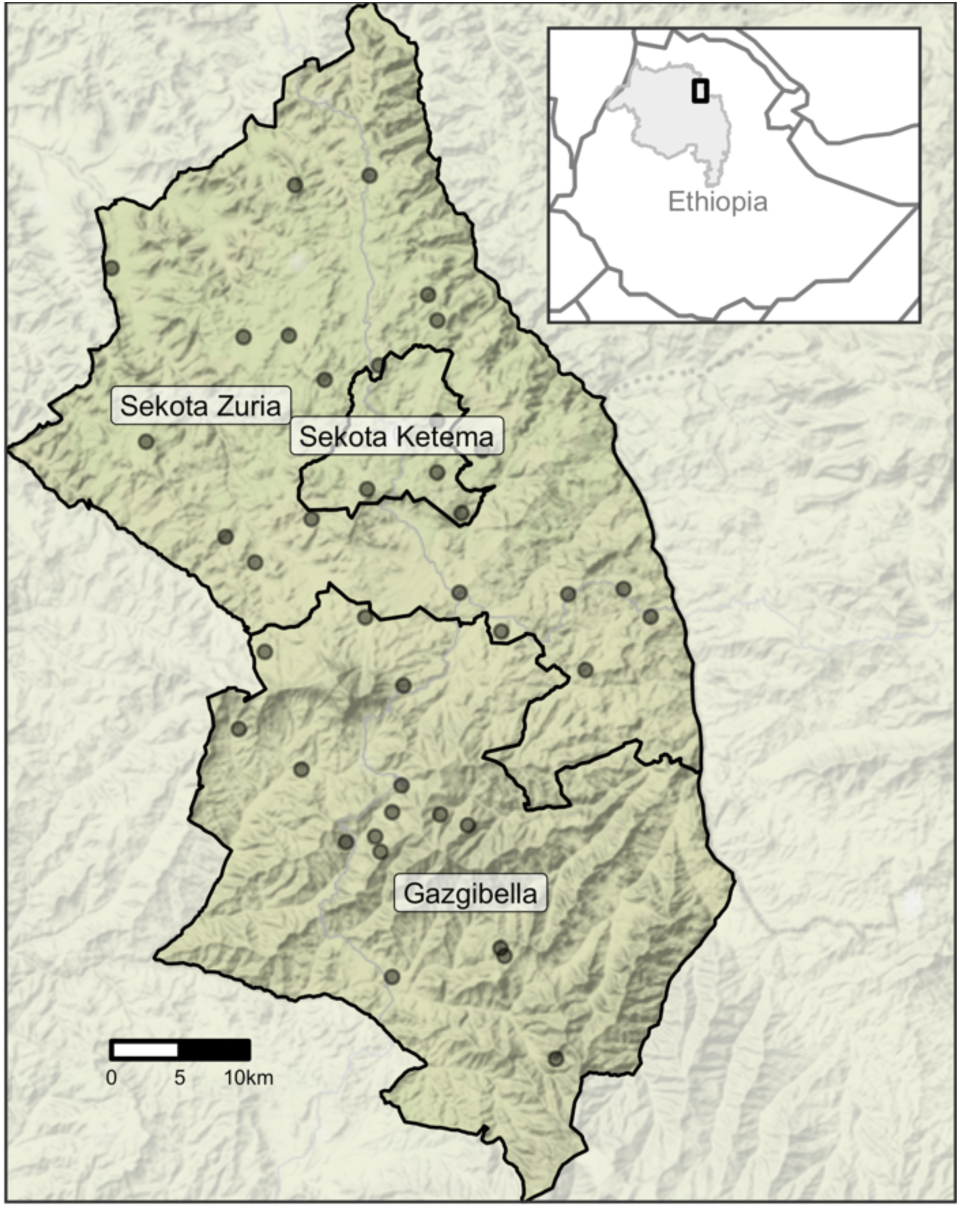
Map of study area. Inset (top right) highlights the Amhara Region (gray shading) of Ethiopia and the study area (black rectangle). Forty communities from three woredas (administrative level 3) in Amhara were included in the WUHA trial.

Approximately thirty children from two age groups (0-5 years old and 6-9 years old) were randomly sampled from each community at baseline and follow-up visits. The number of children evaluated differed slightly for each trachoma indicator (**Table S1**). Over the three-year study period, ocular *Ct* infection prevalence, as measured by PCR, increased substantially in both age groups (**Table 1**). Throughout this analysis, clinical disease was defined as diagnosis with either trachoma inflammation - follicular (TF), the presence of five or more follicles on the upper eyelid, or trachoma inflammation - intense (TI), a condition characterized by inflammatory thickening of the upper eyelid (20). Levels of clinical disease fluctuated with time but remained fairly consistent with baseline levels. Seropositivity, defined as antibody response above pre-determined cut-offs for both Pgp3 and CT694 antigens, increased gradually among 0–5-year-olds. Antibodies were not measured among 6–9-year-olds at months 12 and 24 but were similar between study arms at months 0 and 36. Results were similar when seroprevalence was assessed for each antigen separately (**Table S2**).

**Table 1.**
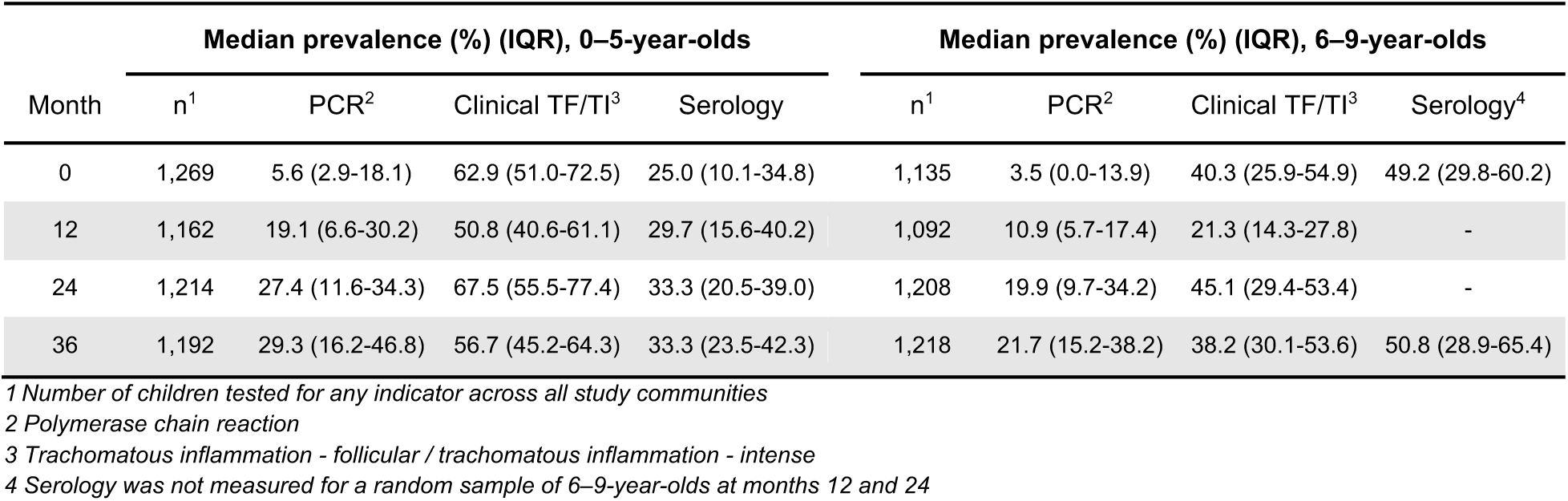
Community-level prevalence of trachoma across 40 study communities by indicator, age group and month of follow-up visit.

| Month | Median prevalence (%) (IQR), 0–5-year-olds |  |  |  | Median prevalence (%) (IQR), 6–9-year-olds |  |  |  |
| --- | --- | --- | --- | --- | --- | --- | --- | --- |
|  | n<sup>1</sup> | PCR<sup>2</sup> | Clinical TF/TI<sup>3</sup> | Serology | n<sup>1</sup> | PCR<sup>2</sup> | Clinical TF/TI<sup>3</sup> | Serology<sup>4</sup> |
| 0 | 1,269 | 5.6 (2.9-18.1) | 62.9 (51.0-72.5) | 25.0 (10.1-34.8) | 1,135 | 3.5 (0.0-13.9) | 40.3 (25.9-54.9) | 49.2 (29.8-60.2) |
| 12 | 1,162 | 19.1 (6.6-30.2) | 50.8 (40.6-61.1) | 29.7 (15.6-40.2) | 1,092 | 10.9 (5.7-17.4) | 21.3 (14.3-27.8) | - |
| 24 | 1,214 | 27.4 (11.6-34.3) | 67.5 (55.5-77.4) | 33.3 (20.5-39.0) | 1,208 | 19.9 (9.7-34.2) | 45.1 (29.4-53.4) | - |
| 36 | 1,192 | 29.3 (16.2-46.8) | 56.7 (45.2-64.3) | 33.3 (23.5-42.3) | 1,218 | 21.7 (15.2-38.2) | 38.2 (30.1-53.6) | 50.8 (28.9-65.4) |

<sup>1</sup> Number of children tested for any indicator across all study communities
<sup>2</sup> Polymerase chain reaction
<sup>3</sup> Trachomatous inflammation - follicular / trachomatous inflammation - intense
<sup>4</sup> Serology was not measured for a random sample of 6–9-year-olds at months 12 and 24

Active ocular infection was more common in the western and northern regions of the study area (**Figure 2A**); seroprevalence and clinical disease were similarly distributed in space (**Figure S1A, Figure S2A**). Based on empirical variograms (**Figure 2B**) and Moran’s I (**Figure 2C**), there was weak spatial structure in community-level Ct PCR prevalence that increased slightly over the study period; serology and clinical indicators also did not display clear spatial structure over the study area (**Figure S1B-C, Figure S2B-C**).

**Figure 2.**
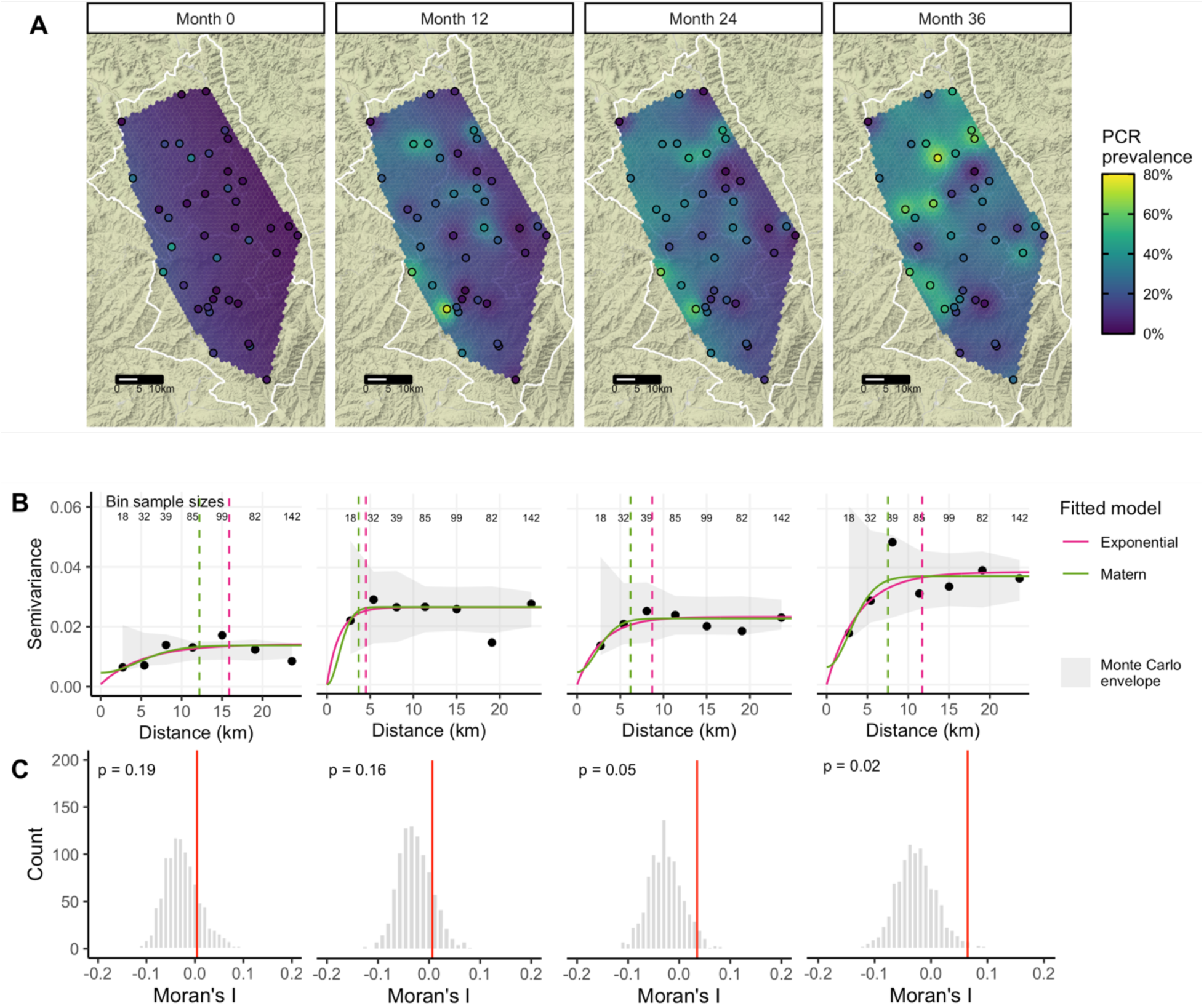
(A) Predicted surface, (B) variograms, and (C) Moran’s I for PCR-confirmed ocular *C. trachomatis* infection prevalence among 0–5-year-olds at each study month. Maps display prevalence for 40 study communities at each follow-up visit, spatially interpolated over the convex hull using kriging. Variograms capture similarity between community-level prevalence measurements as a function of distance between community pairs (in km), with smaller semivariance values representing increased similarity. Exponential (magenta) and Matérn (green) models were fit to each empirical variogram, and the effective range (dashed vertical line) is defined as the distance at which the fitted model reaches 95% of the sill. The Monte Carlo envelope (gray shading) displays pointwise 95% coverage of 1000 permutations, representing a null distribution. Moran’s I was calculated over 1000 permutations (gray bars, with observed value represented by red line), and a permutation-based p-value was calculated.

### Comparisons between serological, clinical, and molecular trachoma indicators

Seroprevalence demonstrated a stronger rank-preserving relationship, as measured by the Spearman correlation, with contemporaneous PCR prevalence than clinical disease for both age groups (**Figure 3A-B**). At baseline, immediately following seven years of MDA, the correlations between trachoma indicators were more pronounced among younger children, potentially reflecting lower transmission in the presence of MDA and saturation in seroprevalence due to durable antibody responses among older children. In a longitudinal cohort nested within the study, children who were seropositive at any survey were very likely to be seropositive one year later (**Figure S4**). Similar saturation dynamics may be at play for clinical trachoma, which has been shown to resolve slowly among children (21). By month 36, when infections were higher across the study area (**Table 1**), correlations between trachoma indicators were similar across age groups (**Figure 3A-B**). Rank-preserving relationships between indicators at each time point and month 36 PCR prevalence were stronger for more proximate measurements, and this increase was more pronounced for PCR compared to clinical trachoma or serology (**Figure 3C**).

**Figure 3.**
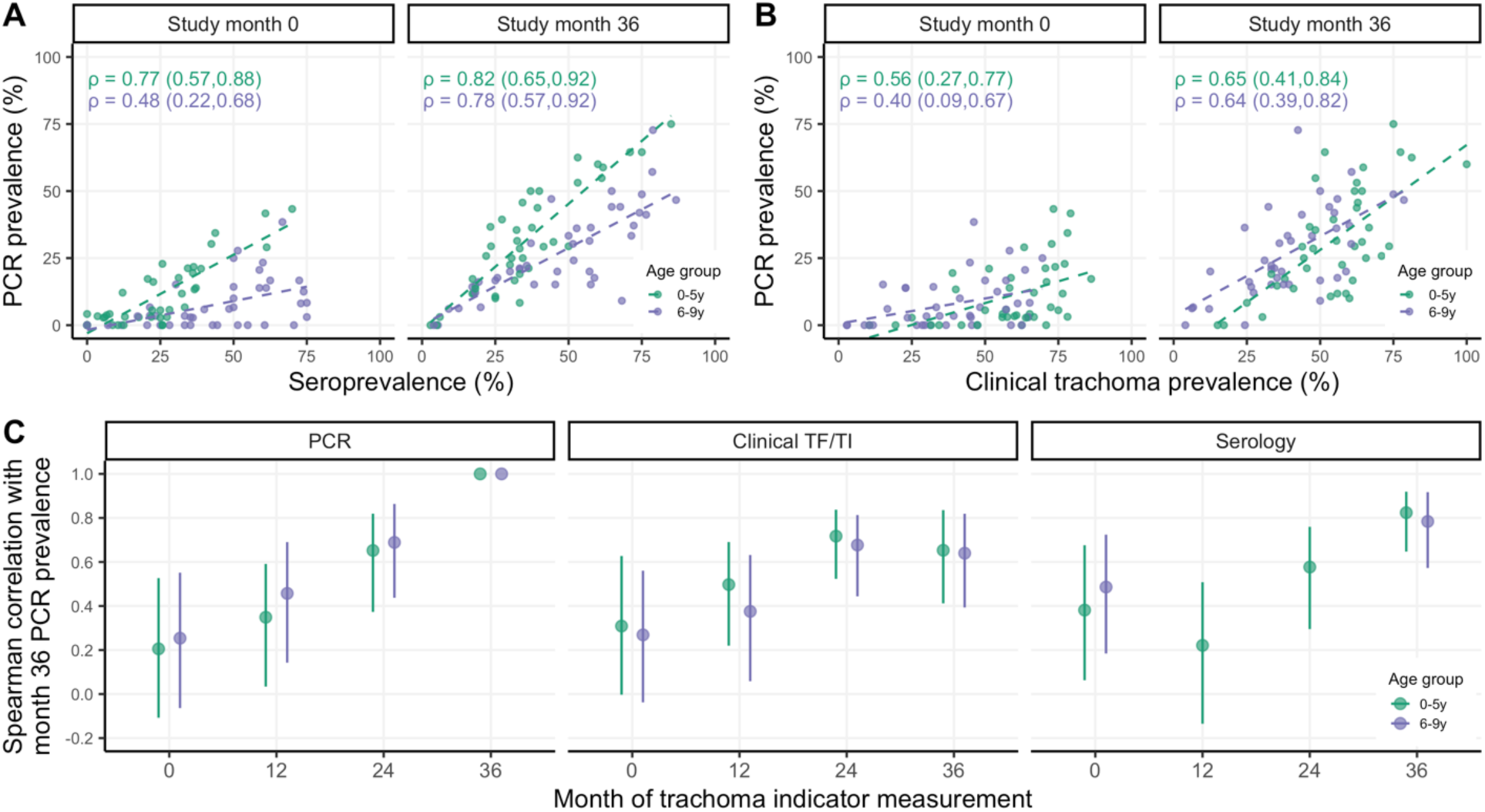
Correlations between trachoma indicators by age group and over time. Panels display Spearman rank correlations between (A) community-level seroprevalence and PCR prevalence at study months 0 and 36, (B) clinical trachoma prevalence and PCR prevalence at months 0 and 36, and (C) PCR prevalence at month 36 and trachoma indicators measured at earlier months across 40 study communities. Correlations are shown separately for 0–5-year-olds (green) and 6–9-year-olds (purple), and 95% confidence intervals were estimated from 1000 bootstrap samples. Serology data was not collected for a random sample of 6–9-year-olds at months 12 and 24.

### Concurrent and forward prediction of PCR prevalence

We predicted community-level infection prevalence using a range of model specifications and conducted spatial 10-fold cross-validation (CV) with 15×15 km blocks (22) to assess predictive performance using CV R^2^ and root-mean-square-error (RMSE) (details in **Materials and Methods**). **Figure 4** presents results for models predicting PCR prevalence at month 36. “Concurrent” predictions utilized trachoma indicators measured at month 36 and/or geospatial variables measured over the preceding year (2018), while “forward” predictions used covariates measured 12, 24, or 36 months in the past. Seroprevalence was the single strongest concurrent predictor of month 36 community-level PCR prevalence (CV R^2^: 0.75, 95% confidence interval (CI): 0.58-0.85, CV RMSE: 0.10), substantially outperforming clinical trachoma prevalence (CV R^2^: 0.37, 95% CI: 0.08-0.56, CV RMSE: 0.16) (**Figure 4**). When predicting 12 months into the future, all trachoma indicators performed moderately well, but predictive performance declined for longer time horizons across all model specifications. No model that we assessed had a CV R^2^ significantly different from 0 (equivalent to an intercept-only or mean-only model) when predicting PCR prevalence 24 months or more into the future.

**Figure 4.**
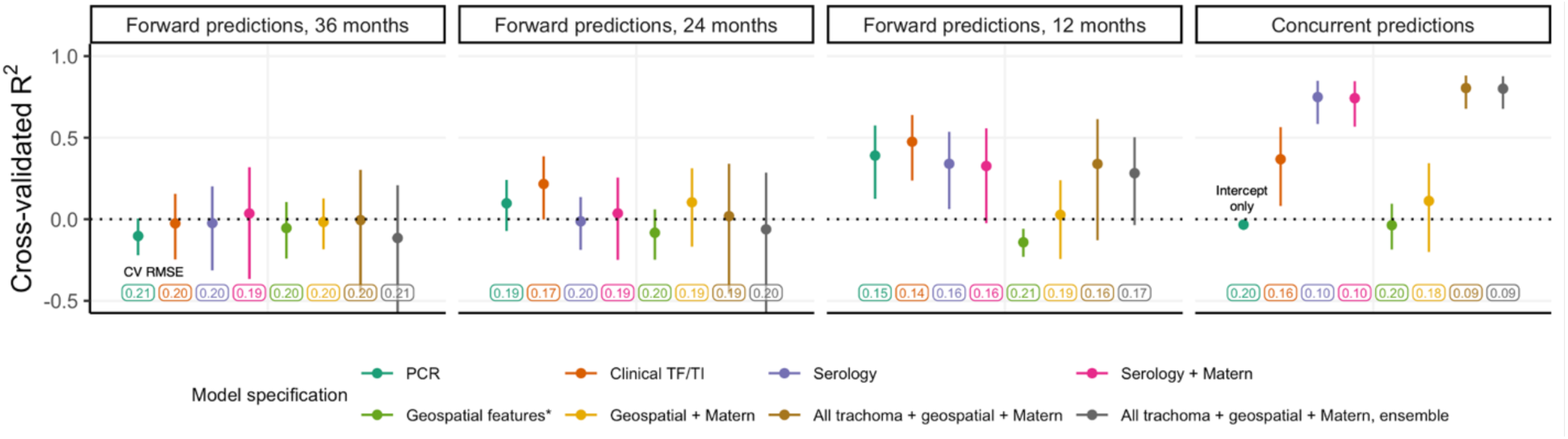
Cross-validated R^2^ for models predicting month 36 community-level PCR prevalence among 0–5-year-olds. Cross-validated coefficient of determination (R^2^), 95% influence-function-based confidence interval, and cross-validated root-mean-square error (RMSE, text label) are shown for each model specification. Logistic regression was used for all models with the exception of the stacked ensemble (gray). Blocks of size 15×15km were used for 10-fold spatial cross-validation.

As anticipated by the weak spatial dependence in PCR prevalence (**Figure 2**), incorporation of a Gaussian process with a Matérn covariance function did not improve predictions. In addition, LASSO-selected geospatial features (night light radiance and daily precipitation averaged over the preceding 12 months) (**Figure S5**) and a stacked ensemble approach leveraging five base models did not meaningfully improve CV R^2^ or CV RMSE compared to simpler models. Results were similar for models predicting PCR prevalence at each time point and pooled over all time points (**Figure S6**).

### Efficient identification of high-burden communities

A complementary task to prediction is identifying communities with the highest infection burden, defined here as the number of *Ct* infections among 0–5-year-olds at a given time. To address variability in sample size, the number of *Ct* infections in each community was scaled to represent a sample of 30 individuals. At month 36, 80% of *Ct* infections were concentrated in just over half of the communities (23/40), and ordering communities by cross-validated concurrent predictions using seroprevalence identified infections more efficiently (i.e. in fewer communities, 25/40) than ordering them by predictions using clinical trachoma (27/40) (**Figure 5**). Performance declined when using predictors measured 12 months in the past, and communities ranked by predictors measured 24 and 36 months in the past could not identify high-burden communities based on PCR infections at month 36 better than chance. The distinction between models was greater at month 0 when 80% of *Ct* infections were concentrated in just the top 15 of 40 (38%) of communities (**Figure S9**).

**Figure 5.**
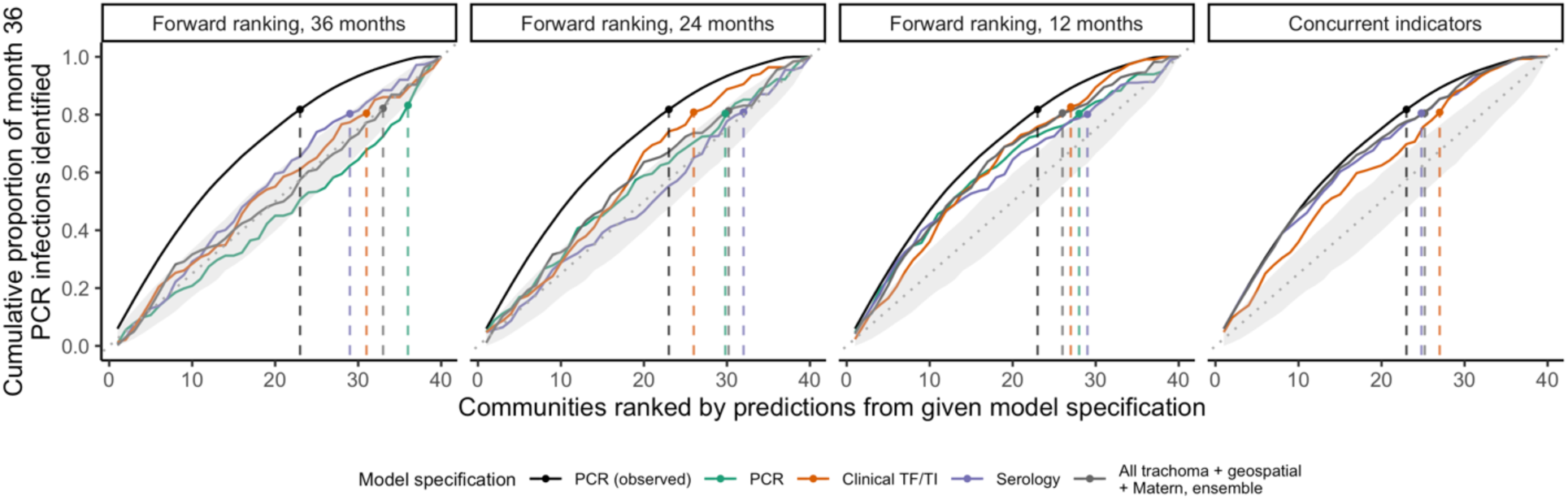
Cumulative proportion of *C. trachomatis* infections at month 36 identified by concurrent and forward prediction models. Dashed lines indicate the point at which the cumulative proportion of identified *Ct* infections at month 36, scaled to represent a sample of 30 individuals per community, surpassed 80%. The black line in each facet represents the optimal ordering of scaled PCR infections at month 36. To simulate a null distribution, we estimated the cumulative proportion of infections identified for 1000 random orderings of the 40 communities and plotted the 95% pointwise envelope (gray shading). For concurrent and 24-month-forward predictions, models using serology only and PCR only, respectively, performed equally well to a model using all trachoma indicators, geospatial features, a Matérn covariance, and ensemble machine learning; vertical lines were offset slightly for visibility.

## DISCUSSION

We conducted a comprehensive study of repeated cross-sectional measurements of clinical trachoma, PCR-positive ocular *Ct* infections, and serological responses to *Ct* antigens over three years in 40 communities in the hyperendemic Amhara region of Ethiopia. In the absence of MDA during the study, active *Ct* infections surged and became increasingly dispersed across study communities. Based on empirical variograms and Moran’s I, we observed weak evidence for global spatial clustering in trachoma indicators over the study region. Seroprevalence among children 0-5 years old aligned closely with PCR prevalence measured at the same time, highlighting the potential for serosurveillance as a monitoring tool that corresponds well with levels of active infection and is potentially easier to measure (23). Predictive performance of all models declined with increasing temporal lag between outcome and predictor measurements. In this setting, remotely sensed demographic, socioeconomic, and environmental geospatial layers, a spatial Gaussian process with Matérn covariance, and stacked ensemble machine learning did not meaningfully improve predictive performance compared to models using only trachoma indicators.

Identifying potential future trachoma hotspots is notoriously challenging and sometimes termed “chasing ghosts” by trachoma programs (17). Our results underscore the difficulty of predicting community-level *Ct* infection prevalence even a year into the future, at least in the context of increasing transmission in the absence of MDA. Furthermore, our “forward prediction” models were trained on infection outcomes from the desired prediction time point and thus were potentially more optimistic than true “forecasting” models trained solely on historical data. Prior efforts to forecast district-level TF (18) and village-level PCR prevalence (17) have explored mechanistic and statistical models and observed modest performance, with one investigation concluding that models with the highest uncertainty resulted in the best predictive performance (17). It remains unclear why future prediction of trachoma presents such a difficult challenge, though likely contributing factors include the stochasticity of rare events especially in near-elimination settings (24), biological unknowns in the complex natural history of trachoma (25), and the extended duration between survey measurements (often 6 months or greater). Models for other neglected tropical diseases have achieved some success in future prediction at the sub-district level, though often capitalizing on larger datasets. For example, a recent study developed models with over 80% accuracy for prediction of *Schistosoma mansoni* persistent hotspots (defined as failure of a village to reduce infection prevalence and/or intensity by specific thresholds) up to two years in the future in the context of decreasing prevalence (26). In a setting with fairly stable transmission, a sub-district-level study for visceral leishmaniasis reported 85.7% coverage of four-month-ahead 25-75% prediction intervals for case counts (27).

Our investigation builds upon an existing body of work characterizing the dynamics between clinical, serological, and molecular trachoma indicators. Reports at the district, village, and individual level have established that relatively high levels of clinical trachoma or ocular infections tend to correspond to higher seroprevalence and/or seroconversion rates (14, 28–31); post-elimination settings have been of particular interest, with populations often displaying little to no antibody response (15, 32–37). Our findings align with earlier studies that showed clinical trachoma is more strongly correlated with infection prevalence in populations with ongoing transmission compared to populations in which transmission has been suppressed by MDA (38–40); also in agreement with prior findings, we observed that TI was slightly, but not significantly, more closely correlated with infection prevalence compared to TF immediately following MDA (**Figure S10**) (41). We additionally found that seroprevalence among children 0-9 years old was more closely aligned with infection prevalence than clinical trachoma in both contexts. Moreover, we found that seroprevalence was more strongly correlated with PCR prevalence among children 0-5 years old compared to children 6-9 years old, especially in the context of recent MDA at month 0. This result supports a focus on children 0-5 years old as a key sentinel population for trachoma serosurveillance.

In general, we did not observe strong evidence of global spatial autocorrelation for trachoma indicators over the study region, though spatial structure in PCR prevalence appeared to increase slightly over the study period. A prior analysis over the entire Amhara region reported evidence of spatial autocorrelation in TF between villages within 25km bands (10), and another study of TF and TI in Southern Sudan detected residual spatial structure between villages at approximately 8 km, after adjusting for age, sex, rainfall, and land cover (42). A larger number of existing studies have characterized spatial autocorrelation at a fairly small scale. Studies using household-level information identified spatial clustering at less than 2 km for bacterial load (6, 9), ocular infection (8, 9), and clinical disease (43). Our ability to detect spatial structure may have been limited by the geographic distribution of the communities, which was determined by the main trial objectives rather than optimized for estimation of spatial model parameters, which often requires points fairly close to one another (44). In our study, only 26 (out of 780) pairs of study communities were within 5 km of one another leading to wide uncertainty at small ranges and hindering our ability to assess fine-scale spatial clustering.

In addition to rainfall and land cover, studies have reported associations between clinical trachoma and distance to water source (10, 45–47), temperature (7, 46, 48), altitude (46, 48– 51), markers of socioeconomic status (7, 10, 45, 47, 51, 52), and markers of personal or household hygiene, such as facial cleanliness (7, 10, 45, 47, 52–59). Fewer studies have examined *Ct* infections identified by PCR, but associations reported were generally similar (52, 59, 60). Using LASSO to down-select geospatial features, we included night light radiance (often a proxy for socioeconomic activity (61)) and precipitation in prediction models. However, these features were unable to predict infection prevalence better than an intercept-only model. Predictive power of geospatial variables may have been limited by relative homogeneity across the study area, and the relatively small number of communities likely limited the predictive performance of all models.

Finally, our analysis focused on a hyperendemic region with increasing trachoma transmission in the absence of MDA and may not generalize to lower transmission settings. Ethiopia’s Amhara region presents a particularly stubborn elimination challenge, as seven consecutive years of MDA were unable to sustain control before the start of this study. It is unclear whether prediction would be more or less challenging in the context of low transmission; we may expect more predictability in a “steady state” environment, but stochasticity is also a defining characteristic of near-elimination disease dynamics (24). As an additional sensitivity analysis, we included survey month as a covariate to assess potential benefits of repeated sampling in the context of changing transmission and found only a modest improvement in predictive performance (**Figure S11**).

### Conclusions

Serological markers among children 0-5 years old may be well-suited for community-level trachoma monitoring given their objectivity, durability, relative ease of collection, and strong correlation with ocular *Ct* infection prevalence. While seroprevalence and clinical trachoma were both correlated with infection prevalence in the midst of high transmission in the absence of MDA, only seroprevalence was strongly associated with community-level infections in the context of suppressed transmission directly following MDA. Accurate, future prediction of community-level *Ct* infection prevalence in settings with unstable transmission remains an open challenge.

## MATERIALS AND METHODS

### Data collection

This work was designed as a secondary analysis of data from the WASH Upgrades for Health in Amhara (WUHA) community-randomized trial, one of the trials in the Sanitation, Water, and Instruction in Face-Washing for Trachoma (SWIFT) (<u>NCT02754583</u>) series. Details of study methodology and implementation are described in the published protocol (19). WUHA was conducted in the Gazgibella, Sekota Zuria (i.e. Sekota) and Sekota Ketema (i.e. Sekota town) woredas of the Wag Hemra Zone in Amhara, Ethiopia. Forty communities were randomized in a 1:1 ratio to receive a comprehensive Water, Sanitation, and Hygiene (WASH) package at baseline or at completion of the study. Mass administration of azithromycin occurred for seven consecutive years (May 2009 to June 2015, with supplemental administration in October 2014) prior to the start of the study but was suspended in all study communities for the duration of the WUHA trial.

Trachoma indicators were measured in each study community at baseline and three annual monitoring visits. Approximately one month prior to each monitoring visit, a census was taken to enumerate individuals living in each study community. The baseline census was conducted in December 2015. At each visit, thirty individuals in three age groups (0-5, 6-9, 10+) were randomly selected from each community for monitoring; this analysis focused on children aged 0-9 years old. Per the trial design, not all trachoma indicators were measured in all age groups at each time point; only children 0-5 years old were tested for clinical, serological, and molecular outcomes at all visits. At the end of WUHA, no difference in the primary endpoint of community-level ocular *Ct* infection among 0–5-year-olds was observed between intervention arms [manuscript under review]. As a result, we combined information across arms for this analysis.

### Measurement and definition of trachoma indicators

We analyzed age-group-specific community-level prevalence of three trachoma indicators: clinical disease, active ocular *Ct* infection detected by polymerase chain reaction (PCR), and IgG response to Pgp3 and CT694 antigens.

Trained trachoma graders used a pair of 2.5X loupes and a flashlight to assess the everted right superior tarsal conjunctiva for the presence of trachomatous inflammation - follicular (TF) or trachomatous inflammation - intense (TI) according to the WHO grading system (62). An individual was considered positive for clinical trachoma if either TF or TI was detected.

Conjunctival swabs were collected and tested in the study laboratory at the Amhara Public Health Institute in Bahir Dar, Ethiopia with the Abbott RealTime assay (automated Abbott m2000 System), which is highly sensitive and specific for *Ct* (63, 64). Groups of five samples, stratified by community and age group, were pooled for testing, and community-level *Ct* infection prevalence was estimated from pooled results using a maximum likelihood approach (65). Certain pools were selected for individual-level PCR testing based on pooled prevalence and other characteristics.

To measure antibody response, field staff lanced the index finger of each individual and collected blood onto TropBio filter paper. Samples were tested at the US Centers for Disease Control on a multiplex bead assay on the Luminex platform for antibodies to two recombinant antigens (Pgp3, CT694) that measure previous exposure to *C. trachomatis* (14, 15, 66). Seropositivity thresholds were defined as median fluorescence intensity minus background (MFI-bg) of 1113 for Pgp3 and 337 for CT694 using an ROC cutoff from reference samples (37). Individuals who were seropositive with respect to both antigens were considered seropositive for the main analysis; descriptive results were similar when considering either antigen separately (**Table S2, Figure S3**).Descriptive analysis of trachoma indicators. Spearman rank correlation coefficients were calculated for pairwise combinations of trachoma indicators by age group and follow-up visit. Correlations were also calculated between PCR prevalence at month 36 and serological, molecular, and clinical prevalence at each preceding time point to observe changes in correlation with increasing temporal lag between measurements. 95% confidence intervals were estimated from 1000 bootstrap samples.

### Descriptive spatial analysis

Administrative boundaries for Ethiopia were downloaded from the Humanitarian Data Exchange (67). Spatially interpolated maps for each trachoma indicator at each time point were generated using a simple kriging model including latitude, longitude, and a Matérn covariance. We estimated empirical variograms after removing linear spatial trends for distances up to 33.3 km (half of the maximum distance between any two study communities) and fit exponential and Matérn models; for stability, we required bins to contain ten or more pairs of communities. The effective, or practical, range was defined as the distance at which the fitted model reached 95% of the sill. We compared the observed variograms to a 95% pointwise envelope based on 1000 Monte Carlo simulations; for each simulation, prevalence residuals were permuted while holding coordinates fixed and the empirical variogram was recalculated (68). We also calculated Moran’s I, a measure of global spatial autocorrelation, over 1000 permutations of the prevalence values and estimated a p-value based on permutations resulting in a Moran’s I greater than or equal to the observed value.

### Predictive model selection

Prediction models were limited to children 0-5 years old due to availability of all trachoma indicators for this age range at all time points. We developed several candidate models using baseline data only, with the analysis team masked to any future measurements. A wide range of publicly available environmental (69–73), demographic (74), and socioeconomic (75–77) variables were explored based on prior associations with trachoma or other infectious diseases (Table S3). When possible, features were extracted and aggregated using Google Earth Engine (78), and means were used for spatial and temporal aggregation unless otherwise specified in Table S3. All features were aggregated to a grid resolution of 2.5 arc minutes (approximately 4.5 km at the median latitude of the study area) based on the lowest resolution dataset (TerraClimate) and reprojected to WGS84. Each community was assigned to the grid cell containing its household-weighted geographic centroid, defined as the median latitude and longitude across all households in the community.

Models were built using predictor variables measured over the same (“concurrent”) and prior (“forward predictions”) time periods. Time-varying features were summarized based on calendar year, with 2015 data considered “concurrent” with month 0 trachoma indicators and so on. Time-varying features were first aggregated by month and then summarized based on recency relative to the time of monitoring (e.g. last 1 month or December of the calendar year, last 2 months, up to 12 months). To reduce collinearity, we evaluated pairwise Pearson correlation coefficients between temporal summaries of the same variable and dropped the summary over fewer months for pairs with correlation over 0.9.

During preliminary model development with baseline data, we observed that a large number of predictor variables led to overfitting and unstable model performance due to the relatively small number of communities. As a result, logistic LASSO regression was used to identify a restricted set of geospatial features to include in the final prediction models. Night light radiance and daily precipitation averaged over the preceding 12 months were selected from a model using concurrently measured predictors and outcomes across all follow-up visits.

Logistic regression models of the following form were used as base prediction models:

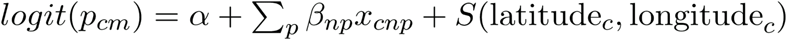

where *p*_*cm*_ represents PCR prevalence for study community *c* at month *m*, α is the model intercept, and *x*_*cn1*_…*x*_*cnp*_ denote covariates with coefficients β measured at time *n*, where *n* = *m* for concurrent predictions and *n = m - k* for predictions *k* months forward. Extended models also included a Gaussian process with Matérn covariance function (79) to capture residual spatial structure, represented by the *S* function dependent on latitude and longitude of the community.

As an extension of our prediction models, we also explored stacked ensemble machine learning, also known as stacked regression (80) or stacked generalization (81). Stacked ensembles combine predictions from multiple ‘Level 0’ models using a ‘Level 1’ model, also called the superlearner or metalearner (82). Ensembles are theoretically guaranteed to perform as well as or better than any single member of their library (80, 82). Our ‘Level 0’ learners included logistic regression, generalized additive models (83), random forest (84), extreme gradient boosting (85), and multivariate adaptive regression splines (86). This set of models, including parametric, semi-parametric, and tree-based methods, was selected to ensure diversity in approach; outcome specification also varied (e.g. binomial, quasibinomial, continuous) based on requirements of the learner. Logistic regression with a Matérn covariance was used as ‘Level 1’ superlearner for the baseline analysis; different superlearner models, including logistic regression without a Matérn covariance and non-negative linear least squares with and without normalized (convex combination) coefficients, resulted in similar predictive performance (**Figure S7**).

### Predictive model assessment

We conducted 10-fold cross-validation to assess predictive performance. Spatial autocorrelation can violate the independence assumption between training and validation sets in cross-validation and lead to overly optimistic estimates of predictive power (22, 87). Therefore, we partitioned the study area into 12 15×15km blocks, each containing 1-8 spatially proximate communities. Communities in the same block were assigned to the same validation set, with some sets consisting of more than one block. This approach decreases spatial dependence between training and validation sets in the same fold and simulates prediction in a new, but geographically proximate, area. We observed consistent results in sensitivity analyses using leave-one-out cross-validation, random cross-validation folds, and spatial blocks of 5×5 km and 20×20 km (**Figure S8**), perhaps reflecting the weak spatial autocorrelation observed in this dataset (**Figure 2**). Predictive performance was assessed using cross-validated root-mean-square-error (RMSE) and R^2^ (88), where R^2^ was calculated as:

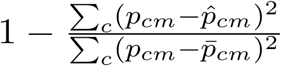

95% confidence intervals for R^2^ were estimated using the influence function (89, 90). Communities received equal weight in all validation metrics.

## Data Availability

Code is currently available on Github; de-identified data will be posted when available.

https://github.com/ctedijanto/swift-spatial-prediction

## Acknowledgements and funding sources

We would like to thank the WUHA study participants and field team without whom this research would not be possible. This work was supported by the National Institute of Allergy and Infectious Diseases (R03 AI147128 to BFA) and the National Eye Institute (U10 EY023939 to JDK). This work was also made possible in part by an Unrestricted Grant from Research to Prevent Blindness. We would also like to thank Abbott for its donation of the m2000 RealTime molecular diagnostics system and consumables.

## Data and availability

The pre-specified statistical analysis plan will be made available on Open Science Framework (https://osf.io). The main R packages used for this analysis were *automap* (variograms) (91), *rgee* (Google Earth Engine) (92), *glmnet* (feature selection) (93), *spaMM* (regression with spatial Gaussian process) (94), *sl3* (stacked ensemble) (95), and *blockCV* (spatial cross-validation) (96). De-identified data and code to replicate this work will be made available on Github (https://github.com/ctedijanto/swift-spatial-prediction). All analysis was conducted in R Version 4.0.2 (“Taking Off Again”) (97).

## Author contributions

Conceptualization: CT, BFA

Data curation: CT, JDK

Formal analysis: CT

Funding acquisition: BFA, JDK

Investigation: CT, BFA

Methodology: CT, HJWS, BFA

Project administration: CT

Resources: SG, DLM, SDN

Software: CT

Supervision: TML, JDK, BFA

Validation: CT

Visualization: CT

Writing - original draft preparation: CT, BFA

Writing - review & editing: All authors

