## Supplementary Information for "Predicting future ocular *Chlamydia trachomatis* infection prevalence using serological, clinical, molecular, and geospatial data"

### Contents

**Table S1.** Number of children evaluated across 40 study communities by trachoma indicator, age group and study month.

**Table S2.** Community-level seroprevalence across 40 study communities by antigen, age group, and study month.

**Table S3.** Description and sources of geospatial variables explored for prediction analysis.

**Figure S1.** Maps (A), variograms (B), and Moran's **I** (C) for seroprevalence among 0–5-year-olds at each study month.

**Figure S2.** Maps (A), variograms (B), and Moran's **I** (C) for clinical trachoma prevalence among 0–5-year-olds at each study month.

**Figure S3.** Correlations between PCR prevalence and antigen-specific seroprevalence by age group and over time.

**Figure S4.** Durability of seropositivity for Pgp3 (A) and CT694 (B) in the WUHA nested longitudinal cohort.

**Figure S5.** Spatio-temporal distribution of LASSO-selected geospatial predictor variables.

**Figure S6.** Cross-validated $R^2$ for models predicting community-level PCR prevalence among 0–5-year-olds at month 0 (A), at month 12 (B), at month 24 (C), at month 36 (D), and pooled across all months (E).

**Figure S7.** Cross-validated $R^2$ for stacked ensemble models predicting community-level PCR prevalence at month 36 among 0–5-year-olds using various superlearner models.

**Figure S8.** Cross-validated $R^2$ for models predicting community-level PCR prevalence among 0–5-year-olds at month 36 using random 10-fold cross-validation (A), 10-fold spatial cross validation with 5x5 km blocks (B), 15x15 km blocks (C), and 20x20 km blocks (D), and leave-one-out cross-validation (E).

**Figure S9.** Cumulative proportion of *C. trachomatis* infections at months 0 and 36 identified by concurrent prediction models.

**Figure S10.** Correlations between PCR prevalence and clinical signs of trachoma by age group and over time.

**Figure S11.** Cross-validated $R^2$ for models predicting pooled community-level PCR prevalence among 0–5-year-olds at month 36 with survey month (time) modeled as a linear covariate or Gaussian process.

### Table S1. Number of children evaluated across 40 study communities by trachoma indicator, age group and study month.

| | Number evaluated for indicator, 0–5-year-olds | | | | Number evaluated for indicator, 6–9-year-olds | | | |
|---|---|---|---|---|---|---|---|---|
| Month | Overall | PCR[1] | Clinical TF/TI[2] | Serology | Overall | PCR[1] | Clinical TF/TI[2] | Serology[3] |
| 0 | 1,269 | 1,258 | 1,256 | 1,245 | 1,135 | 1,129 | 1,085 | 1,109 |
| 12 | 1,162 | 1,154 | 1,151 | 1,122 | 1,092 | 1,090 | 1,072 | 0 |
| 24 | 1,214 | 1,210 | 1,206 | 1,200 | 1,208 | 1,206 | 1,204 | 0 |
| 36 | 1,192 | 1,183 | 1,181 | 1,188 | 1,218 | 1,212 | 1,193 | 1,214 |

*1 Polymerase chain reaction*
*2 Trachomatous inflammation - follicular / trachomatous inflammation - intense*
*3 Serology was not measured for a random sample of 6–9-year-olds at months 12 and 24*

### Table S2. Community-level seroprevalence across 40 study communities by antigen, age group, and study month.

| | Median prevalence (%), 0–5-year-olds (IQR) | | | | Median prevalence (%), 6–9-year-olds (IQR)[3] | | | |
|---|---|---|---|---|---|---|---|---|
| Month | n[1] | Serology[2] | Pgp3 | CT694 | n[1] | Serology[2] | Pgp3 | CT694 |
| 0 | 1,245 | 25.0 (10.1-34.8) | 30.4 (13.3-40.7) | 26.5 (10.1-34.9) | 1,109 | 49.2 (29.8-60.2) | 60.0 (39.4-70.6) | 49.2 (32.1-61.5) |
| 12 | 1,122 | 29.7 (15.6-40.2) | 36.0 (21.8-47.5) | 30.7 (15.6-46.8) | 0 | - | - | - |
| 24 | 1,200 | 33.3 (20.5-39.0) | 38.2 (24.0-45.8) | 33.3 (20.5-39.7) | 0 | - | - | - |
| 36 | 1,188 | 33.3 (23.5-42.3) | 40.0 (32.5-52.2) | 33.3 (24.6-42.3) | 1,214 | 50.8 (28.9-65.4) | 58.7 (34.6-77.5) | 50.8 (28.9-65.4) |

*1 Number of children tested for serology*
*2 Seropositive for both Pgp3 and CT694*
*3 Serology was not measured for a random sample of 6–9-year-olds at months 12 and 24*

**Table S3. Description and sources of geospatial variables explored for prediction analysis.**

| Variable | Description | Temporal resolution | Source |
|---|---|---|---|
| ***Environmental variables*** | | | |
| Precipitation | Precipitation (mm) | Daily | CHIRPS (69) |
| Maximum temperature | Maximum temperature (°C) | Monthly | TerraClimate (70) |
| Minimum temperature | Minimum temperature (°C) | Monthly | TerraClimate (70) |
| EVI | Enhanced vegetation index | 16-day period | MODIS (71) |
| Elevation | Average digital elevation (km) | Static, 2000 | SRTM (72) |
| Slope | Derived terrain slope (degrees) | Static, 2000 | SRTM (72) |
| Surface water | Landsat-derived presence/absence of surface water at any time during the month | Monthly | Joint Research Center Global Surface Water (73) |
| ***Demographic variables*** | | | |
| Population | Overall population density | Static, 2018 | HRSL (74) |
| Age distribution | 0-5 years old, proportion of population | Static, 2019 | HRSL (74) |
| Sex distribution | Female, proportion of population | Static, 2019 | HRSL (74) |
| ***Socioeconomic variables*** | | | |
| Night lights | Radiance (nanoWatts/cm2/sr) excluding negative values and pixels based on 5 or fewer cloud-free observations | Monthly | VIIRS (76) |
| Distance to roads | Euclidean distance from center of grid cell to nearest road (km) | Static, 2020 | OpenStreetMap (75) |
| Access to care | Land-based travel time to nearest healthcare facility (minutes), all transport and non-motorized transport only (i.e. walking-only) | Static, 2019 | Malaria Atlas Project (77) |

**Figure S1. Maps (A), variograms (B), and Moran's I (C) for seroprevalence among 0–5-year-olds at each study month.** Maps display prevalence for 40 study communities at each follow-up visit, spatially interpolated over the convex hull using kriging. Variograms capture similarity between community-level prevalence measurements as a function of distance between community pairs (in km), with smaller semivariance values representing increased similarity. Exponential (magenta) and Matérn (green) models were fit to each empirical variogram, and the effective range (dashed vertical line) is defined as the distance at which the fitted model reaches 95% of the sill. The Monte Carlo envelope (gray shading) displays pointwise 95% coverage of 1000 permutations, representing a null distribution. Moran's I was calculated over 1000 permutations (gray bars, with observed value represented by red line), and a permutation-based p-value was calculated.

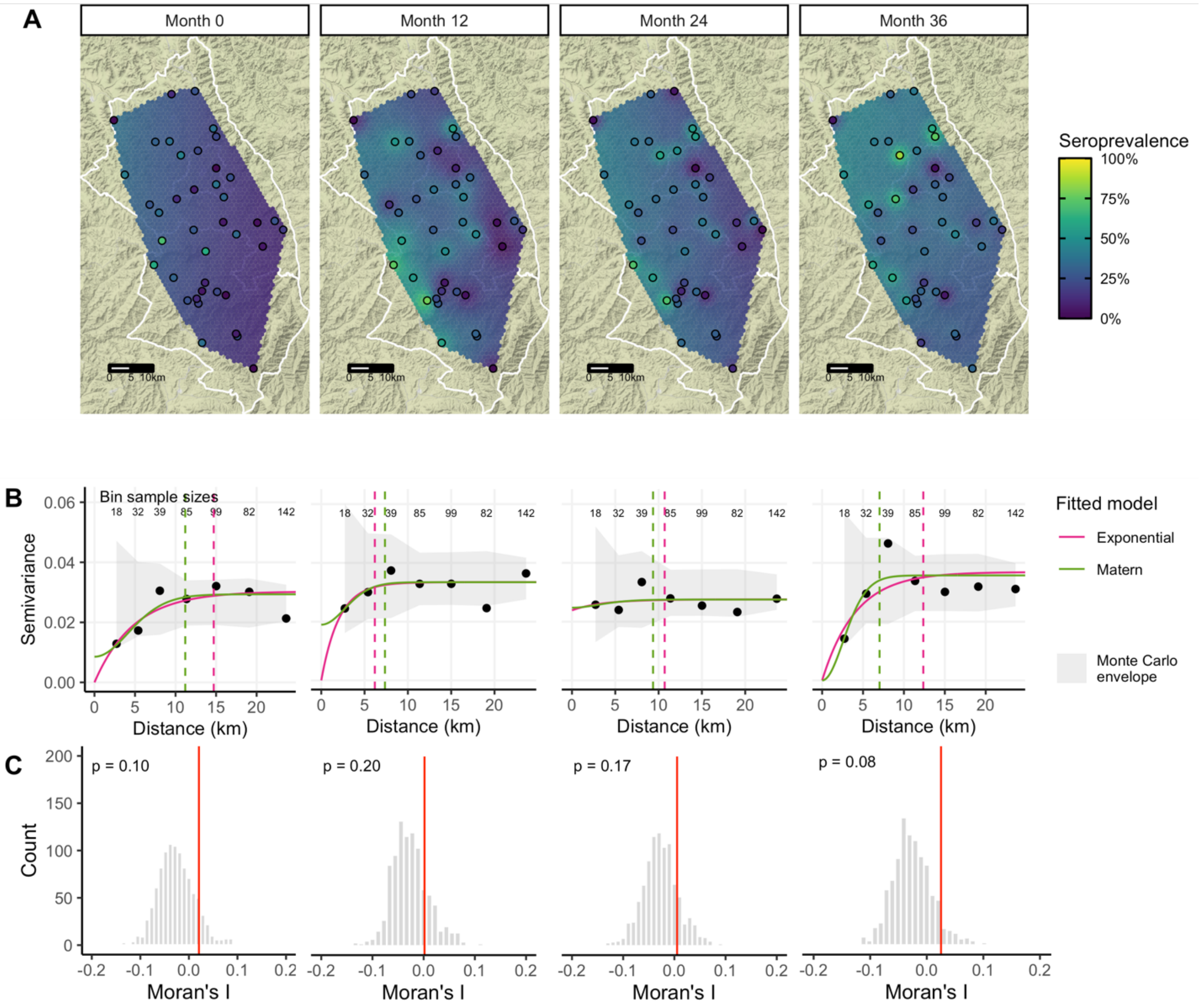

**Figure S2. Maps (A), variograms (B), and Moran's I (C) for clinical trachoma prevalence among 0–5-year-olds at each study month.** Maps display prevalence for 40 study communities at each follow-up visit, spatially interpolated over the convex hull using kriging. Variograms capture similarity between community-level prevalence measurements as a function of distance between community pairs (in km), with smaller semivariance values representing increased similarity. Exponential (magenta) and Matérn (green) models were fit to each empirical variogram, and the effective range (dashed vertical line) is defined as the distance at which the fitted model reaches 95% of the sill. The Monte Carlo envelope (gray shading) displays pointwise 95% coverage of 1000 permutations, representing a null distribution. Moran's I was calculated over 1000 permutations (gray bars, with observed value represented by red line), and a permutation-based p-value was calculated.

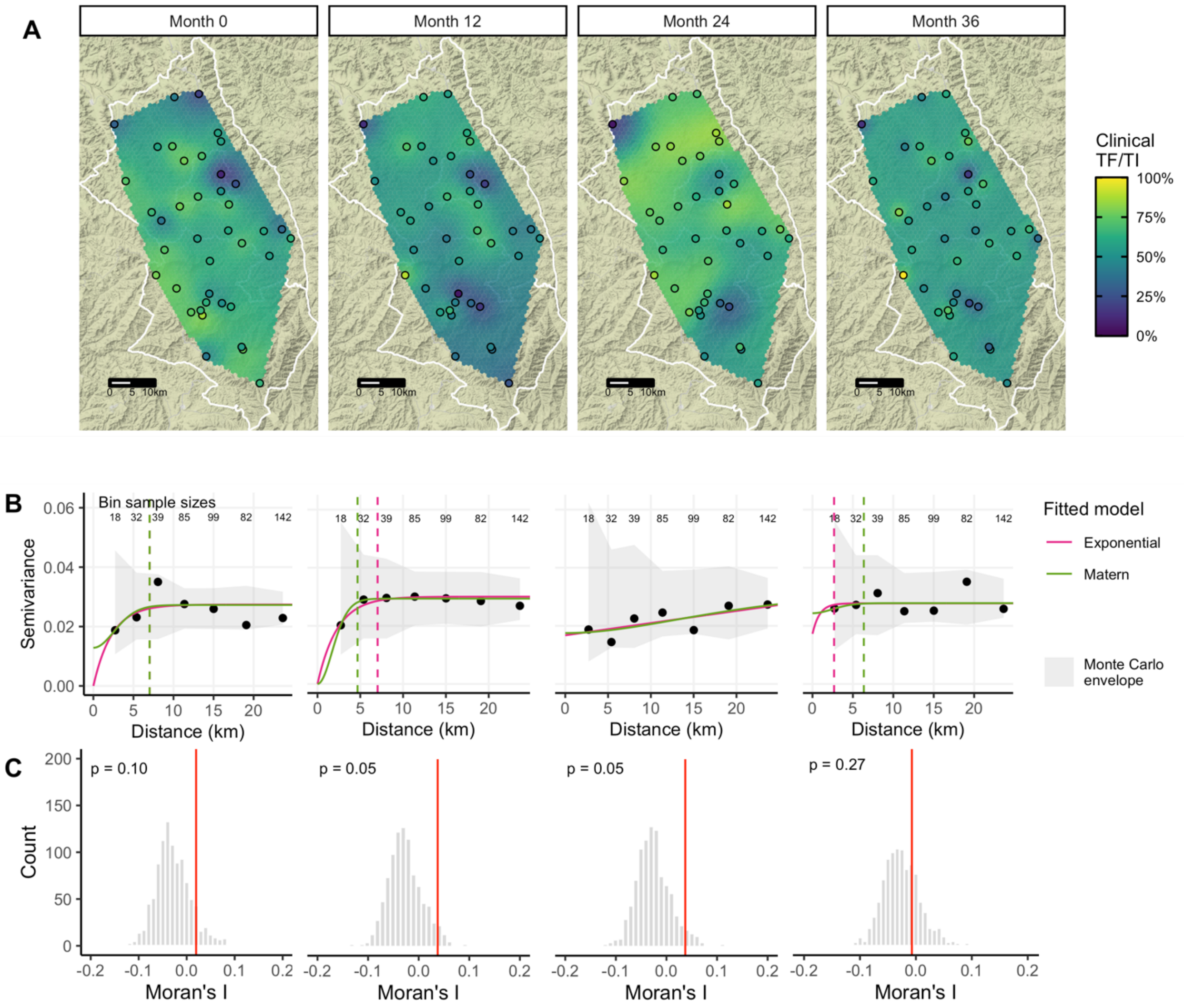

**Figure S3. Correlations between PCR prevalence and antigen-specific seroprevalence by age group and over time.** Panels display Spearman rank correlations between (A) community-level Pgp3 prevalence and PCR prevalence at months 0 and 36, (B) CT694 prevalence and PCR prevalence at months 0 and 36, and (C) PCR prevalence at month 36 and seroprevalence measured at each follow-up visit across 40 study communities. Correlations are shown separately for 0–5-year-olds (green) and 6–9-year-olds (purple) when possible, and 95% confidence intervals were estimated from 1000 bootstrap samples. Serology data was not collected for a random sample of 6–9-year-olds at months 12 and 24.

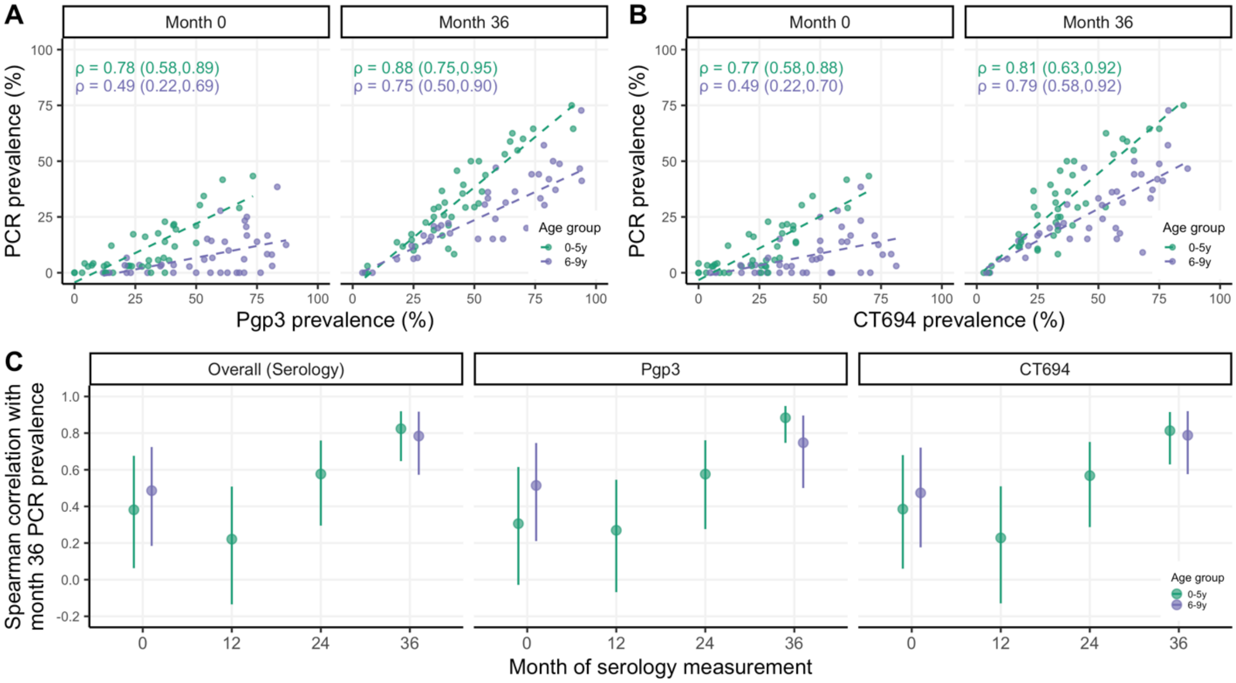

**Figure S4. Durability of seropositivity for Pgp3 (A) and CT694 (B) in the WUHA nested longitudinal cohort.** The random sample of 0–5-year-olds selected at month 0 from each study community was followed longitudinally, and clinical, serological, and molecular samples were collected as described in the **Materials and Methods**. IgG antibody response, represented by the Median Fluorescence Intensity (MFI) minus background (bg), at month *x+12* is plotted among children who were seropositive at month *x* and had serology measured at month *x+12*. Antibody responses at month *x+12* are stratified by individual PCR status. Dotted horizontal lines represent seropositivity cutoffs for each antigen.

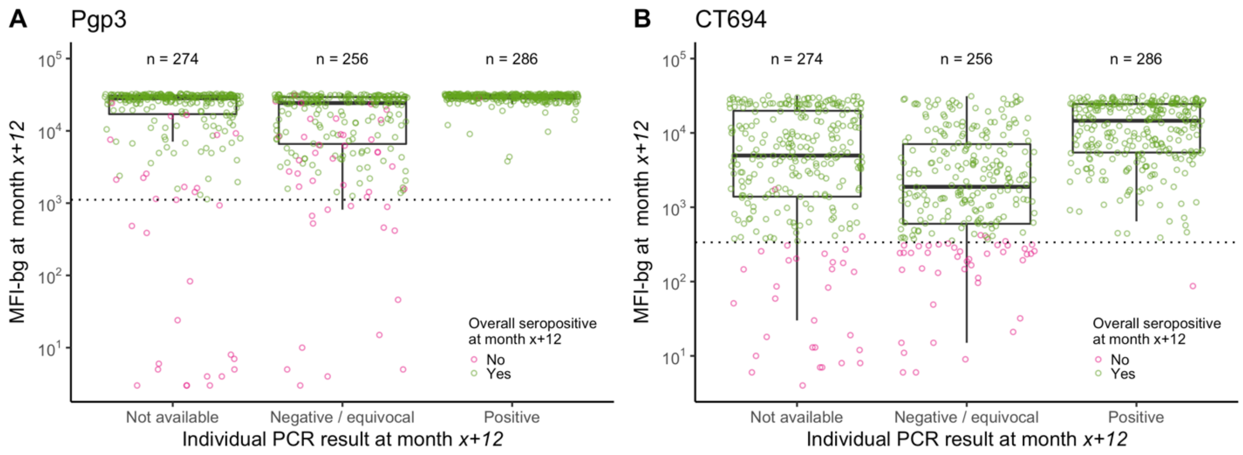

**Figure S5. Spatio-temporal distribution of LASSO-selected geospatial predictor variables.** Variables were estimated for 240 grid cells of 2.5 x 2.5 arc minutes (approximately 20 $km^2$ at the median latitude of the study area). (A) Daily precipitation and (B) monthly night light radiance averaged over the year were included in the final set of prediction models.

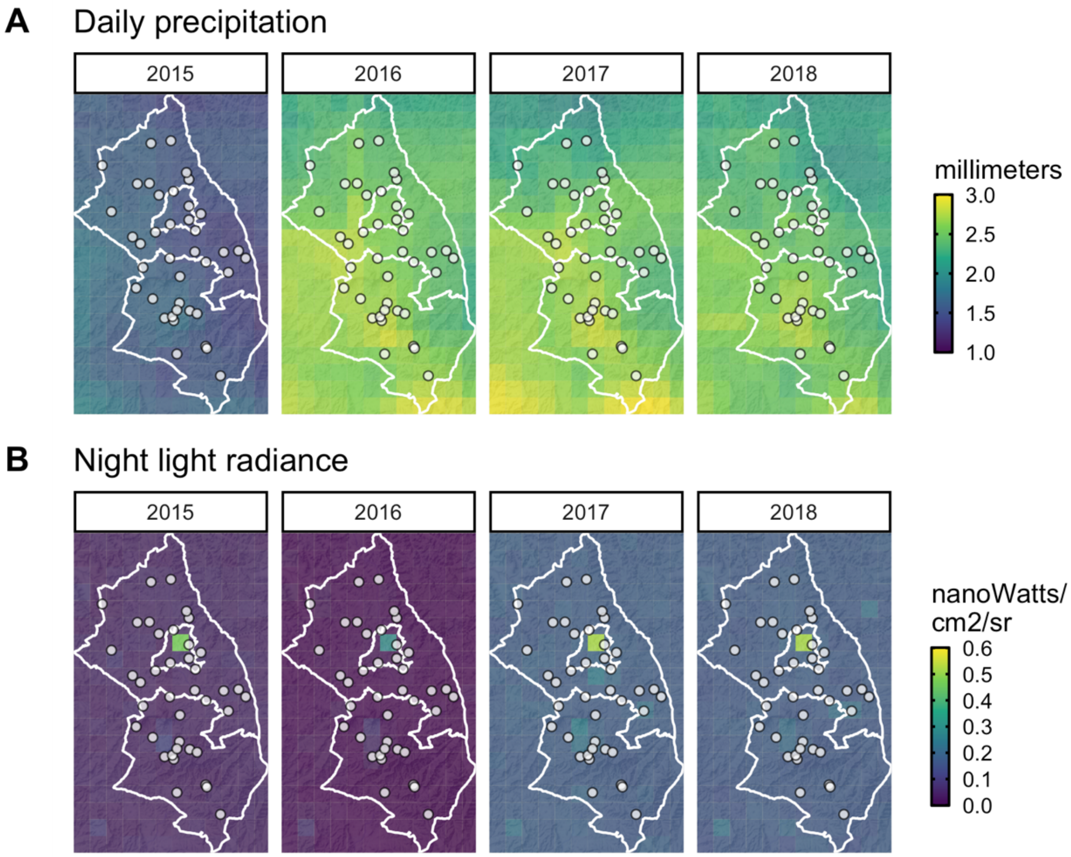

**Figure S6. Cross-validated $R^2$ for models predicting community-level PCR prevalence among 0–5-year-olds at month 0 (A), at month 12 (B), at month 24 (C), at month 36 (D), and pooled across all months (E).** Cross-validated $R^2$ (coefficient of determination), 95% influence-function-based confidence interval, and cross-validated root-mean-square error (RMSE, text label) are shown for each model specification. Blocks of size 15x15km were used for 10-fold spatial cross-validation. (D) is equivalent to **Figure 4** in the main text and is included here for comparison.

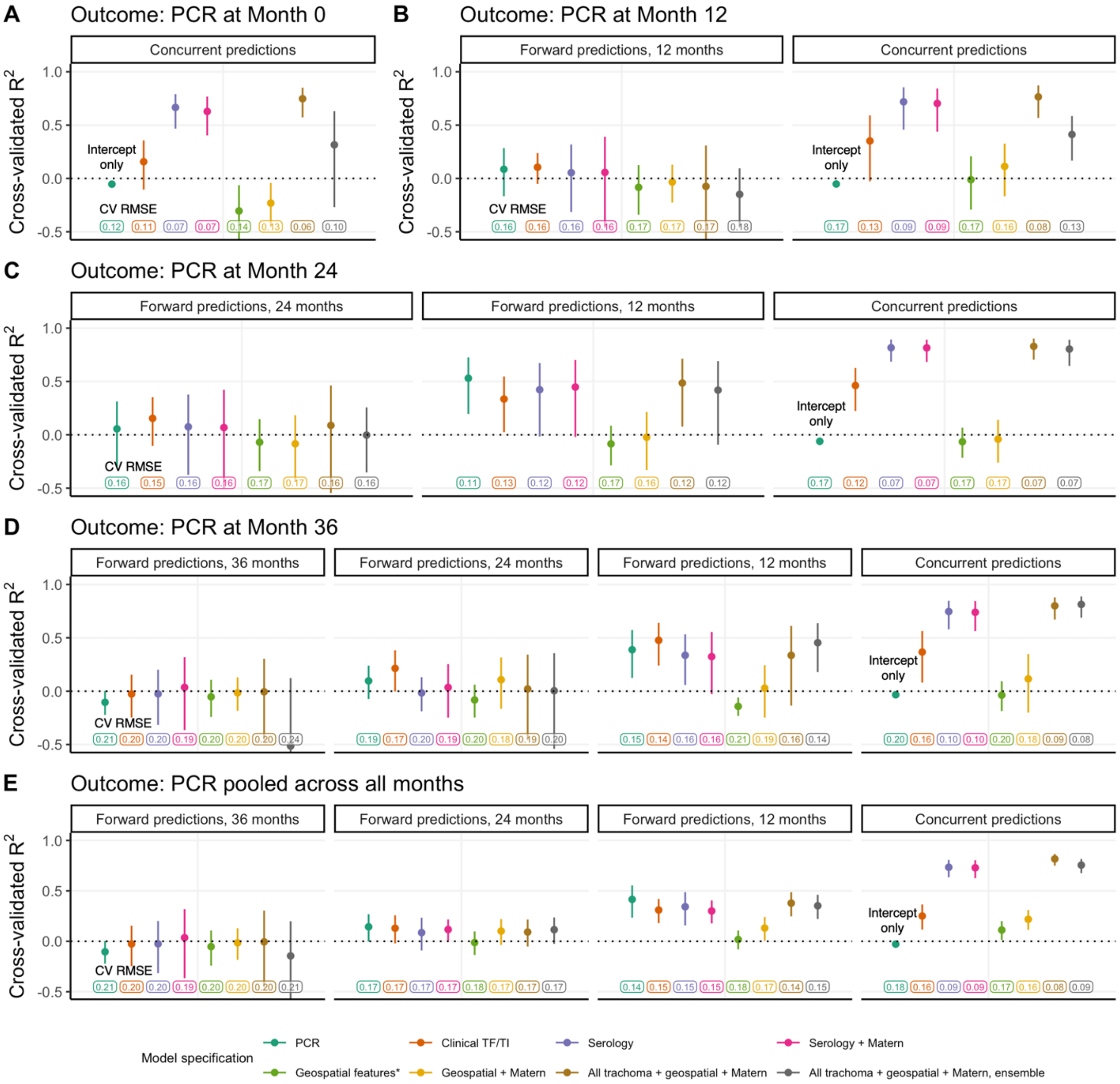


*Includes 12-month precipitation and night light radiance as selected by LASSO

**Figure S7. Cross-validated $R^2$ for stacked ensemble models predicting community-level PCR prevalence at month 36 among 0–5-year-olds using various superlearner models.** Cross-validated $R^2$ (coefficient of determination), 95% influence-function-based confidence interval, and cross-validated root-mean-square error (RMSE, text label) are shown for each model specification. Blocks of size 15x15km were used for 10-fold spatial cross-validation.

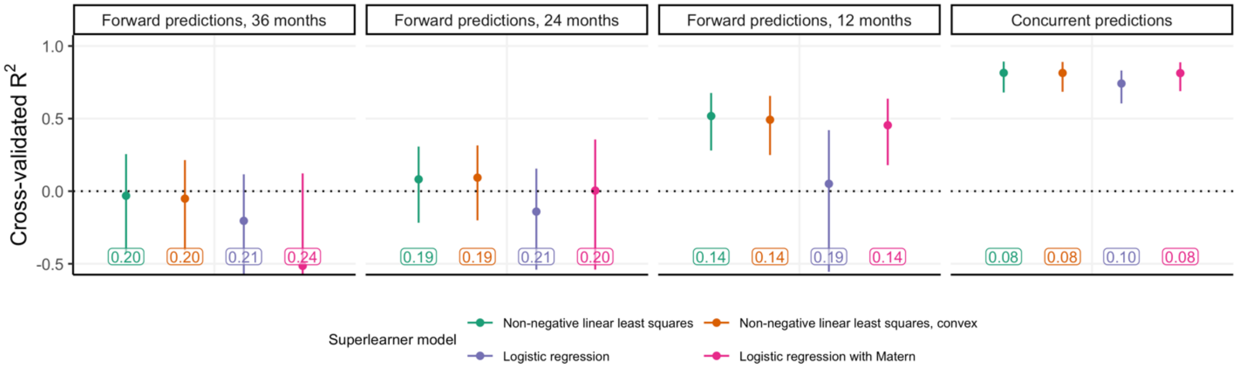

**Figure S8. Cross-validated $R^2$ for models predicting community-level PCR prevalence among 0–5-year-olds at month 36 using random 10-fold cross-validation (A), 10-fold spatial cross validation with 5x5 km blocks (B), 15x15 km blocks (C), and 20x20 km blocks (D), and leave-one-out cross-validation (E).** Cross-validated $R^2$ (coefficient of determination), 95% influence-function-based confidence interval, and cross-validated root-mean-square error (RMSE, text label) are shown for each model specification. (C) is equivalent to **Figure 4** in the main text and is included here for comparison.

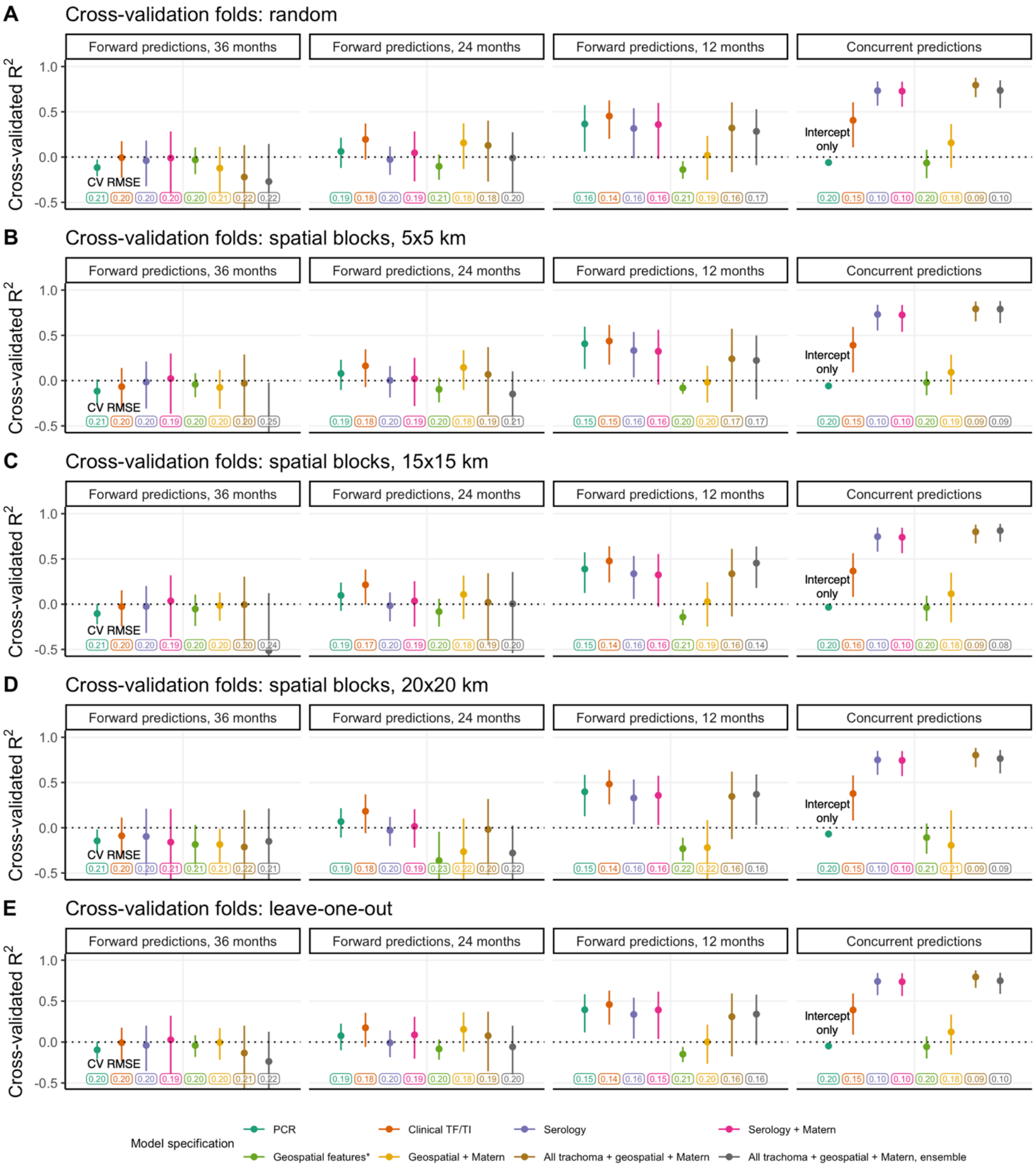


*Includes 12-month precipitation and night light radiance as selected by LASSO

**Figure S9. Cumulative proportion of *C. trachomatis* infections at months 0 and 36 identified by concurrent prediction models.** The black line in each facet represents the optimal ordering of scaled PCR infections at each respective month. Dashed lines indicate the point at which the cumulative proportion of infections, scaled to represent a sample of 30 individuals per community, surpassed 80%. To simulate a null distribution, we estimated the cumulative proportion of infections identified for 1000 random orderings of the 40 communities and plotted the 95% pointwise envelope (gray shading). At month 36, a model using only serology performed equally well to a model using all trachoma indicators, geospatial features, a Matérn covariance, and ensemble machine learning; vertical lines were offset slightly for visibility.

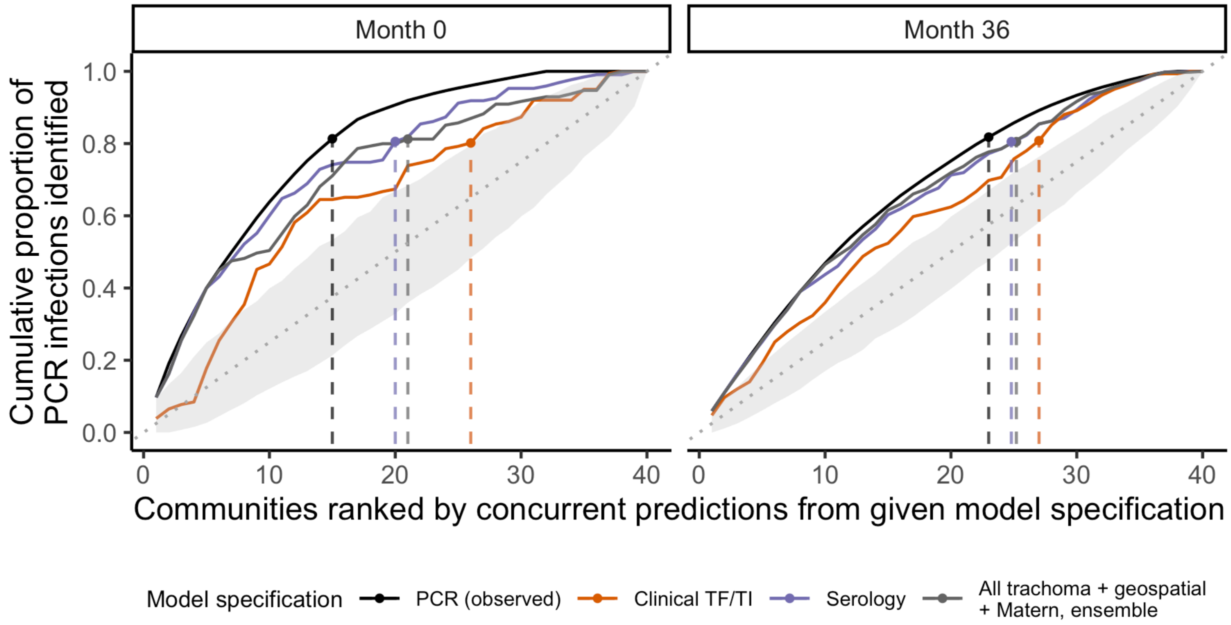

**Figure S10. Correlations between PCR prevalence and clinical signs of trachoma by age group and over time.** Panels display Spearman rank correlations between (A) community-level TF prevalence and PCR prevalence at months 0 and 36, (B) TI prevalence and PCR prevalence at months 0 and 36, and (C) PCR prevalence at month 36 and clinical signs measured at each follow-up visit across 40 study communities. TF prevalence included any child diagnosed with TF, regardless of TI status, and vice versa. Correlations are shown separately for 0–5-year-olds (green) and 6–9-year-olds (purple), and 95% confidence intervals were estimated from 1000 bootstrap samples.

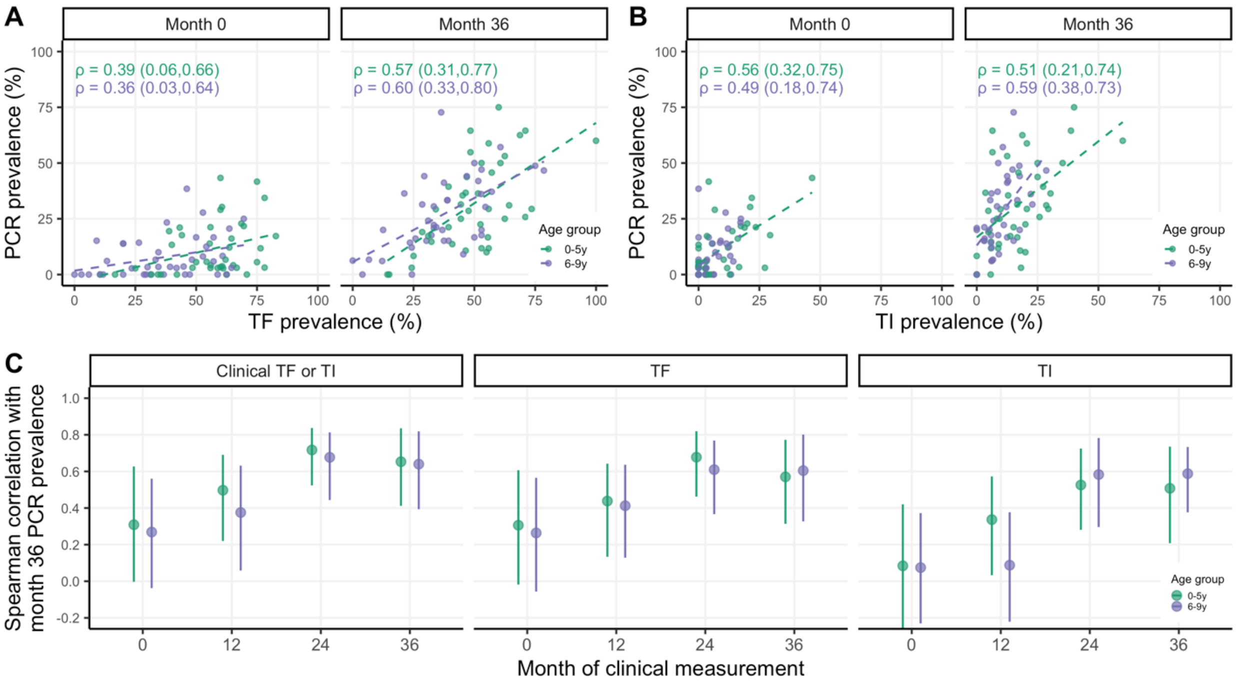

**Figure S11. Cross-validated $R^2$ for models predicting pooled community-level PCR prevalence among 0–5-year-olds at month 36 with survey month (time) modeled as a linear covariate or Gaussian process.** Cross-validated $R^2$ (coefficient of determination), 95% influence-function-based confidence interval, and cross-validated root-mean-square error (RMSE, text label) are shown for each model specification. Blocks of size 15x15km were used for 10-fold spatial cross-validation. For predictions 36 months ahead, time could not be explicitly modeled as a linear covariate as all outcomes were measured at month 36.

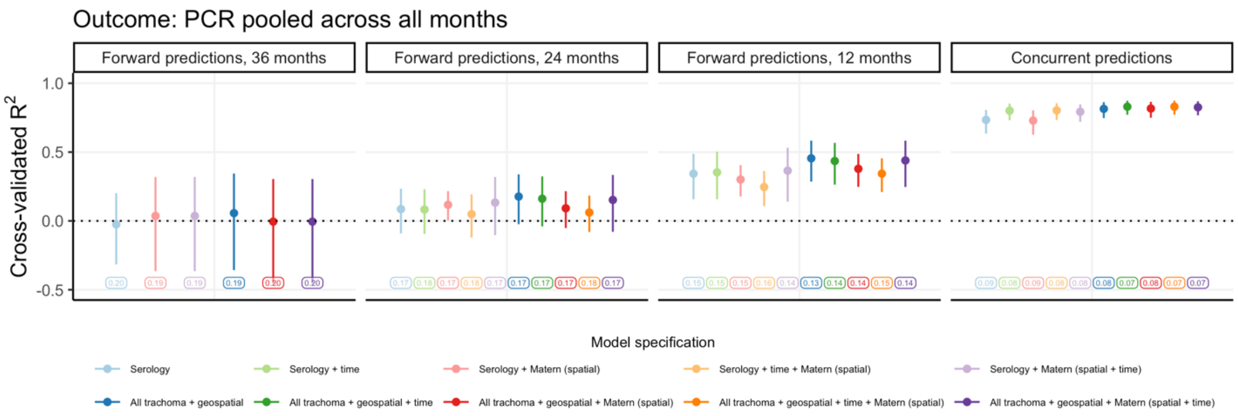


*Includes 12-month precipitation and night light radiance as selected by LASSO
